# Assessing empirical thresholds for investigation in people referred on a symptomatic colorectal cancer pathway: a cohort study utilising faecal immunochemical and blood tests in England

**DOI:** 10.1101/2023.03.29.23287919

**Authors:** Colin J Crooks, Ayan Banerjea, James Jones, Caroline Chapman, Simon Oliver, Joe West, David J Humes

## Abstract

**Objective:** To quantify risk of colorectal cancer (CRC) at empirical FIT cut offs, across age, haemoglobin and platelet strata in current diagnostic pathways.

**Design:** Cohort study of all people who were referred on a symptomatic CRC diagnosis pathway from primary care with a FIT test in Nottingham, UK between November 2017 and 2021 with 1-year follow-up for cancer and death. Heat maps showed the cumulative 1-year CRC risk using Kaplan-Meier estimates. We estimated the number of investigations that could potentially be re-purposed if a threshold of ≥3% 1-year risk of CRC was instigated.

**Results:** During the study period 514 (1.5%) colorectal cancers were diagnosed following 33694 index FIT tests with available blood tests. Individuals with a FIT ≥10 μg Hb/g faeces had a greater than 3% risk of CRC, except patients under the age of 40 years (CRC risk 1.45% (95% CI 0.03-2.86%)). Non-anaemic patients with a FIT <100 μg Hb/g faeces had a CRC risk of less than 3%, except those between the age of 70-85 years (5.26% 95% CI 2.72-7.73%). Using a ≥3% CRC threshold in patients < 55 years calculated using FIT, age and anaemia would allow 160-220 colonoscopies per 10000 FIT tests to be used for other pathways, at the cost of missing 1-2 CRCs.

**Conclusions:** CRC risk varies by FIT, age and anaemia status when fHb levels are below 100 μg Hb/g faeces. Tailored cut offs for investigation on a CRC pathway could reduce the number of investigations needed at a 3% CRC risk threshold.

**What is already known on this topic:** The National Institute for Health and Care Excellence (NICE), the joint Association of Coloproctology of Great Britain & Ireland and the British Society of Gastroenterology guidelines and the Welsh Government recommend a FIT cut off of ≥10 μg Hb/g faeces for investigation of CRC on an urgent pathway based on an expected colorectal cancer risk threshold of 3%. However, empirical evidence of this threshold in practice and the impact of age, anaemia and thrombocytosis upon it is lacking.

**What this study adds:** People who had a FIT test in primary care in Nottingham between 2017 and 2021 had a 1- year risk of colorectal cancer of just 1.5%.

Non-anaemic patients over 70 years old do not meet the 3% threshold set by NICE for urgent investigation until they have a FIT greater than 40 μg Hb/g faeces.

Patients under 40 years of age only meet the 3% threshold for investigation when they have a FIT ≥100 μg Hb/g faeces and are anaemic.

**How this study might affect research, practice or policy:** We estimate that by using a stratified approach to meet the ≥3% risk of CRC threshold that includes FIT, age and anaemia rather than a single cut off for FIT of ≥10 μg Hb/g faeces will allow the optimum number of cancers to be diagnosed for the investigations undertaken.

This study assessed existing empirical categorisations of FIT, age and anaemia. Ideally, further optimisation and validation of pathways could be achieved by deriving cut offs and strata using continuous modelling of FIT, age and blood test results.

The balance of investigations required, cancers diagnosed and missed is crucial to consider when attempting to optimise diagnostic accuracy and health service provision in the real world. Consensus among all stakeholders needs to be reached on the threshold (risk of CRC) at which investigation should be triggered, taking all these factors into account.

## Introduction

In the 2015 National Institute for Health and Care Excellence (NICE) guidelines “Suspected Cancer: recognition and referral (NG12)” the stated “risk threshold” for referral or direct access endoscopy was a 3% positive predictive value of cancer i.e. those referred by primary care on a cancer diagnostic pathway should have a risk of the specific cancer of 3% or more to trigger further investigation^1 2^. This threshold was chosen to improve the diagnosis of cancer over a previously used threshold of 5% by targeting those at greatest risk for the most appropriate investigation^1^ but was a pragmatic compromise as patient preferences are for lower cut offs^3^. In colorectal cancer (CRC) the pathway to diagnosis has rapidly evolved over the past 5 years incorporating Faecal Immunochemical Testing (FIT) as the chosen biomarker in the field^1 2 4^. The COVID-19 pandemic has accelerated the use of FIT in symptomatic patients across the United Kingdom in an attempt to manage the massive backlog in endoscopy, yet the implementation has been piecemeal and not evidenced based^5-7^. One of the big unknowns in this space is the actual risk of CRC that occurs at specific cut offs of FIT when used freely in Primary Care, and what impact use of these cut offs will have on diagnostic investigation services, as highlighted in the recent Association of Coloproctology of Great Britain & Ireland and the British Society of Gastroenterology guidelines^4^.

Many guidelines, including those from NICE, and those recently developed jointly by the Association of Coloproctology of Great Britain & Ireland (ACGBI), the British Society of Gastroenterology (BSG) and the Welsh Government have endorsed the use of FIT in symptomatic patients^1 2 4 8^. They recommend the use of FIT to guide clinicians in choosing the most appropriate pathway for symptomatic patients (without a palpable rectal mass) – specifically that patients with a faecal haemoglobin ≥10 μg Hb/g faeces should be selected for referral on an urgent diagnostic pathway whilst other, less urgent, pathways should be considered below this cut off. Unfortunately, the choice of a FIT ≥10 μg Hb/g faeces cut off has been made without the necessary population-based studies with enough people included to assess the impact use of such a cut off has and without acknowledgement that a value of 10 μg Hb/g faeces might not be equivalent across analysers^9^. The cut off of ≥10 μg Hb/g faeces was originally suggested for patients with a low risk of CRC who were being seen in primary care and further research was recommended for those considered at higher risk and the optimum cut off to use^1 10^. It is crucial to balance the risk of CRC against the ability of the health service to investigate and diagnose cancer in an appropriate and timely manner. Other factors will influence the risk of CRC and therefore the interpretation of a FIT result – notably age, evidence of anaemia or inflammation. In addition, the actual risk of CRC with a FIT result of ≥10 μg Hb/g faeces is much lower than 3%^11^. This means that it is likely that many people (i.e. those under 50 years of age, those with normal blood parameters) are being investigated for CRC who have a very small chance of having the disease. This, inevitably, will contribute to diagnostic services being overwhelmed with the associated reduction in capacity and risk of delays in diagnosis for others. This is especially important given the NHS long term plan to increase the proportion of Stage 1 and 2 cancers to 75% by 2028 which will require additional endoscopy capacity to facilitate a reduction in Bowel Cancer Screening Programme Age from 60 to 50 years^12^.

To assess the risk of CRC in people with a FIT test among a population referred via a single diagnostic pathway and quantify the CRC risks at different cut offs of FIT, age groups and by the presence of anaemia or inflammation we have used all available electronic health data associated with FIT referrals over a 4-year period spanning before and during the COVID-19 pandemic. Our aim was to determine the empirical thresholds of CRC risk in a representative population at different FIT cut offs to assess optimal use of FIT in patients with symptoms of CRC.

## Methods

### Nottingham Rapid Colorectal Cancer Diagnosis Pathway (NRCCD)

In Nottingham, a locally commissioned 12-month service evaluation of FIT in urgent CRC pathways allowed “local agreement” of a new pathway designed by all stakeholders (General Practitioners (GPs), secondary care, Clinical Care Commissioning Groups (CCGs), the Bowel Cancer Screening Hub). This incorporated FIT as a triage tool for symptomatic suspected CRC referrals, except rectal bleeding and palpable rectal mass, as described elsewhere ^13-15^. The data from this service evaluation led to a re-design of the local pathways. Following approval and rollout of this pathway in November 2017^7^, GPs were able to request FIT (and blood tests) independently for the investigation of CRC (Appendix I). FIT and FBC were mandated for all CRC referrals, other than rectal bleeding or rectal mass, irrespective of symptoms or age. FIT and blood results were used to prioritise access to urgent investigations based on multiple thresholds and published evidence, continuous local data evaluation and national context was used to guide iterative changes to the pathway as described in supplementary figure 1.

### Study setting

The study was conducted at Nottingham University Hospitals (NUH) NHS Trust, using data for all primary care requested FIT tests for suspected CRC, processed within the Bowel Cancer Screening Hub within pathology services at Nottingham University Hospitals NHS Trust (NUH) among^16^:

- Adults (>= 18 years of age)
- Patients within Nottinghamshire registered at a General Practice that would refer to Nottingham University Hospitals (Nottingham City and South Nottingham Integrated Care Partnerships)
- From 01/Nov/2017 until 31/Nov/2021.

FIT requests and results reporting was electronic. FIT dispatch and return were entirely postal and kits were analysed according to manufacturer’s protocols by our accredited BCSP Hub laboratory. All samples were analysed using an OC-Sensor™ platform (Eiken Chemical Co., Tokyo, Japan) as previously described^15^.

### Exclusion criteria

FIT tests done in patients younger than 18 years old or those who were not registered with a GP in Nottingham City and South Nottingham ICPs or FIT tests conducted outside the study time period.

### Data Management

The variables of interest were extracted and linked by patients’ unique identification numbers using Microsoft SQL Server. The data were then anonymised prior to being accessed by the researchers, so the researchers did not have access to identifiable patient level data and no patient level data left NUH NHS Trust. The anonymous data for analysis were transferred to a separate secure server within NUH that only the investigative team could access for analysis (CC, JW), so no patient data left NUH NHS Trust.

### Outcomes

CRC was defined from linked Infoflex (Civica) data where all cancers diagnosed at NUH NHS Trust are recorded. Fact and date of death was obtained from the NHS personal demographics service and underlying cause of death (coded with ICD-10) from https://www.hed.nhs.uk/Info/.

### Exposures

Each individual had their first recorded FIT test per year (index FIT test) identified and were subsequently linked to all the required datasets within NUH NHS Trust’s Enterprise Data Warehouse. This included age (at date of FIT test) using year of birth, sex (defined as male/female) and blood tests which were extracted for all patients and included haemoglobin/platelets using any result up to one year prior and 14 days following the index FIT test. The closest of these tests to the index FIT test in time were used. Ferritin was not measured frequently enough to be included in this study. Cut offs from the Nottingham pathway and published literature were used to define strata within this study fHb <4 μg Hb/g faeces, 4-9.9 μg Hb/g faeces, 10-19.9 μg Hb/g faeces, 20 – 39.9 μg Hb/g faeces, 40-99.9 μg Hb/g faeces and ≥100 μg Hb/g faeces, along with anaemia (≤130g/l in men; ≤120g/l in women), and abnormal platelet count (≥400 x10^9^/l)^11 15^. We identified all relevant investigations for CRC (colonoscopy, flexible sigmoidoscopy, Computed Tomography colonography) that occurred within 6 months of the index FIT test.

### Statistical analysis

The analyses were carried out using R^17^ within R Studio on NUH NHS Trust devices. We constructed a study population of all included patients with their FIT result, demographics, relevant test results and outcomes (CRC, death (cause)). We described the baseline characteristics and crude risks of outcomes. We described the time from index FIT test to diagnosis and/or death using histograms. Plots of the different continuous variables and the risk of the binary outcomes were assessed visually for linear and non-linear relationships. We assessed the completeness of the data and have classified missing data as missing for this descriptive study. Patients were followed up for up to one year for CRC diagnosis or death. Diagnosis of CRC was stratified by FIT category using the cut offs defined above. We also undertook an analysis with a FIT level of ≥10 μg Hb/g faeces as per the current National guidance^18^. We stratified the analyses by age, presence or absence of anaemia and thrombocytosis. These data were used to produce heat maps showing 1-year cumulative CRC risks using 1 - Kaplan-Meier survival estimate for CRC with 95% confidence intervals, within strata. We quantified the number of people who had relevant investigations for CRC within 6 months of the index FIT test. Finally, we estimated the number of investigations that could potentially be re-purposed if investigation was restricted to groups with a 3% or greater 1-year CRC risk and the number of CRCs that would be missed by such a strategy.

## Results

### Demographics

In total 34435 patients returned 39774 FIT tests, with 37216 FIT tests with one year follow up, after excluding 2558 (6.4%) returning more than one test within a year of the initial test. Only 6% of the population were under the age of 40 years and the number of FIT tests requested was greatest in those aged 55-85 years (Table 1).

**Table 1.** Demographics of the Nottingham FIT cohort (all patients and tests including those missing blood tests, n = 37216).

|  | Number of patients with measurement | Number with repeated FIT tests | Number of tests within 14 days or year prior to FIT test | % missing with results carried within 14 days or prior year | N (%) or median value (IQR) |
| --- | --- | --- | --- | --- | --- |
| **Gender** |  |  |  |  |  |
| Male | 15061 | 1120 |  |  | 16274 (44%) |
| Female | 19374 | 1438 |  |  |  |
| **Age (Years)** |  |  |  |  |  |
| Age 18-40 | 2247 | 74 |  |  | 2278 (6%) |
| Age 40-55 | 7210 | 370 |  |  | 7516 (20%) |
| Age 55-70 | 10991 | 761 |  |  | 11738 (32%) |
| Age 70-85 | 11803 | 1126 |  |  | 12927 (35%) |
| Age >85 | 2598 | 227 |  |  | 2757 (7%) |
| **Test results** |  |  |  |  |  |
| FIT Test | 34435 | 2558 | 37216 | 0 | 4 (4,8) |
| Blood tests |  |  |  |  |  |
| Hb (g/dl) | 30999 |  | 33694 | 9.5 | 131 (118,143) |
| Platelet Count | 30901 |  | 33586 | 9.8 | 268 (223,322) |
| Ferritin | 28182 |  | 30725 | 17.4 | 65 (23,139) |
| **Investigations** |  |  |  |  |  |
| Colonoscopy, CT colonography within 6 months |  |  |  |  |  |
| Colonoscopy | 6540 |  |  |  |  |
| CT colonography | 3103 |  |  |  |  |
| **CRC diagnoses and Deaths within 1 year** |  |  |  |  |  |
| Colorectal cancer | 533 |  |  |  | 1.5 |
| Colorectal cancer deaths | 79 |  |  |  | 0.2 |
| Non colorectal cancer deaths | 1469 |  |  |  | 4.3 |

### Colorectal Cancer diagnoses

During the study period a total 533 (1.5%) CRCs were diagnosed following the index FIT test. In the year following the index FIT test there were 79 deaths from CRC and 1469 (4.3%) deaths from other causes. The largest proportion of CRC were diagnosed in those patients with a FIT > 100 μg Hb/g faeces (Table 2, only showing patients with blood tests available). Median time to diagnosis was 35.8 days (33.7 – 39.8 days), to non-CRC death was 165.6 days (157.6 – 176.1 days) and the distribution of diagnoses and deaths relative to the index FIT test are shown in supplementary results.

**Table 2.**
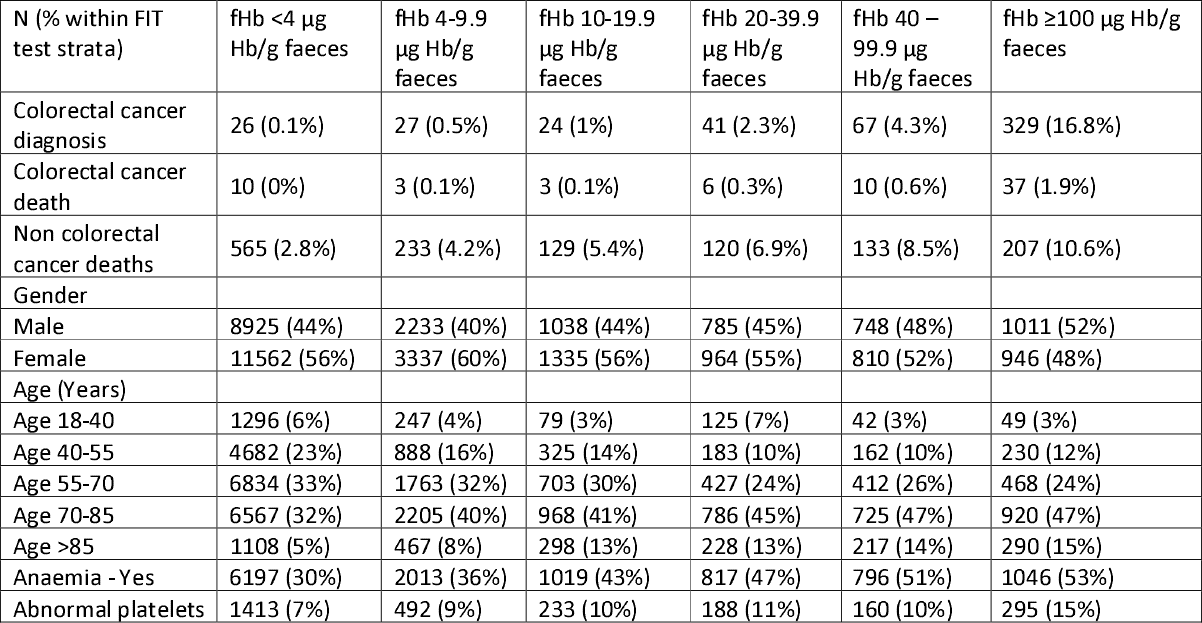
CRC diagnoses and exposures by FIT level in the Nottingham pathway (using only first FIT per year in patients with FIT tests with blood tests, n = 33694, total CRC = 514).

### Missing data

9.7% of patients had no recorded valid haemoglobin measurement, and 9.8% no recorded valid platelet count within one year prior and 14 days post FIT test. The remainder of the results are presented in those with complete data, consisting of 33,694 unique FIT results from 30,999 patients with 514 (1.5%) CRC diagnoses.

### 1-year cumulative CRC risks by age and FIT level

Only 26 (5.1%) cancers were diagnosed in those patients with a FIT <4 μg Hb/g faeces. Most cancers were diagnosed in those patients with a FIT of ≥100 μg Hb/g faeces 329 (64%). At a reported FIT of <10 μg Hb/g faeces 53 cancers were diagnosed compared to 461 with a FIT ≥10 μg Hb/g faeces. Stratifying FIT level by age demonstrated that all patients with a FIT <10 μg Hb/g faeces had a 1-year cumulative CRC risk of less than 3% (Figure 1). All patients with a FIT ≥10 μg Hb/g faeces had a greater than 3% risk of CRC except those under the age of 40 years who had a cumulative CRC risk of 1.45% (95% CI 0.03-2.86%). Stratifying by all our FIT cut offs (figure 2) shows that the 3% threshold for FIT is ≥100 μg Hb/g faeces for patients under 70 years, and ≥40 μg Hb/g faeces for those over 70 years. Using a lower 95% confidence interval bound of the Kaplan Meier estimate would require a FIT cut off of ≥20 μg Hb/g faeces for all patients over 40 years.

**Figure 1.**
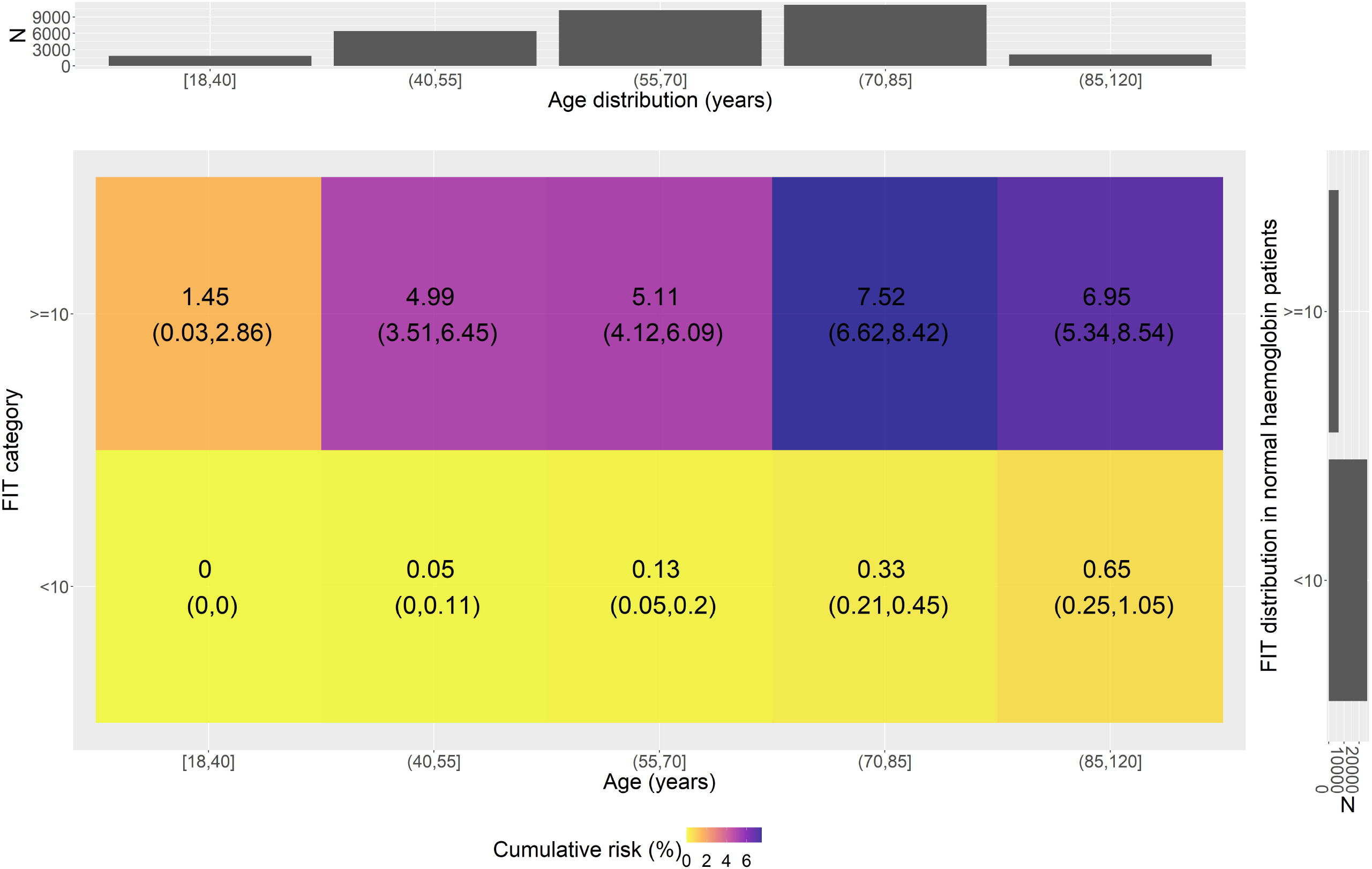
Heat map of CRC diagnoses by FIT level dichotomised at ≥10 μg Hb/g (using only patients with blood tests, n = 33694)

**Figure 2.**
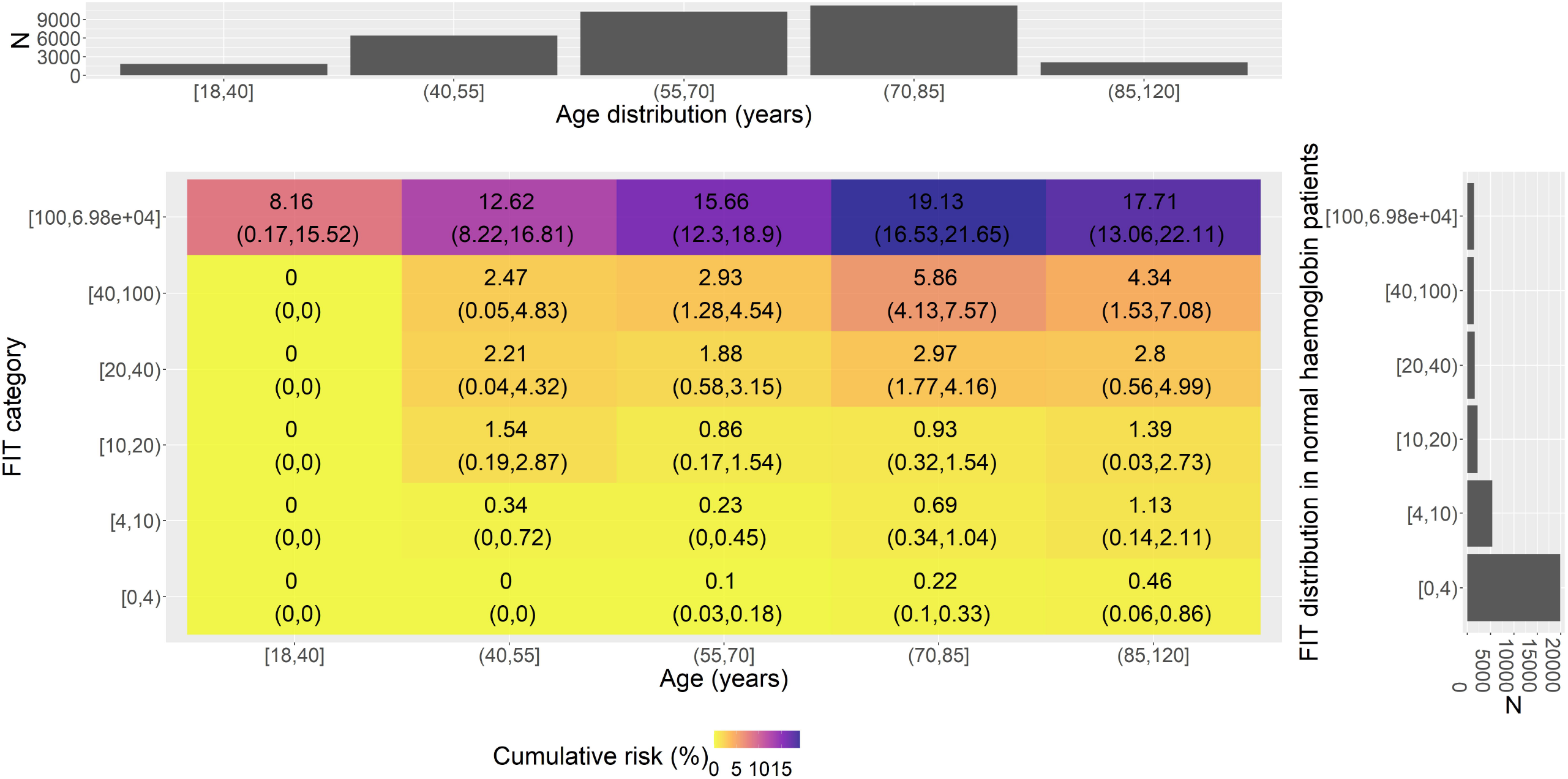
Heat map of CRC diagnoses by FIT level (5 categories) and age group (using only patients with blood tests, n = 33694)

### 1-year cumulative CRC risks by age, anaemia and FIT level

Following stratification, of CRC diagnosis by age, FIT level and anaemia, non-anaemic patients with a FIT <100 μg Hb/g faeces had a cumulative CRC risk of less than 3% (Figure 3) except those between the age of 70-85 years (5.26% 95% CI 2.72-7.73%). Using a lower 95% confidence interval bound of the Kaplan Meier estimate would require a cut off ≥40 μg Hb/g faeces for all non-anaemic patients under 70 years, and ≥20 μg Hb/g faeces for patients over 70 years. In contrast, in patients who had anaemia the 3% threshold was met over 40 years in the FIT 20-40 μg Hb/g faeces category. In anaemic patients under 40 years of age no further cancers were detected in those with a FIT <100 μg Hb/g faeces. Using a lower 95% confidence interval bound of the Kaplan Meier estimate in anaemic patients would require a FIT cut off ≥100 μg Hb/g faeces for all patients under 40 years, and ≥20 μg Hb/g faeces for patients over 40 years.

**Figure 3.**
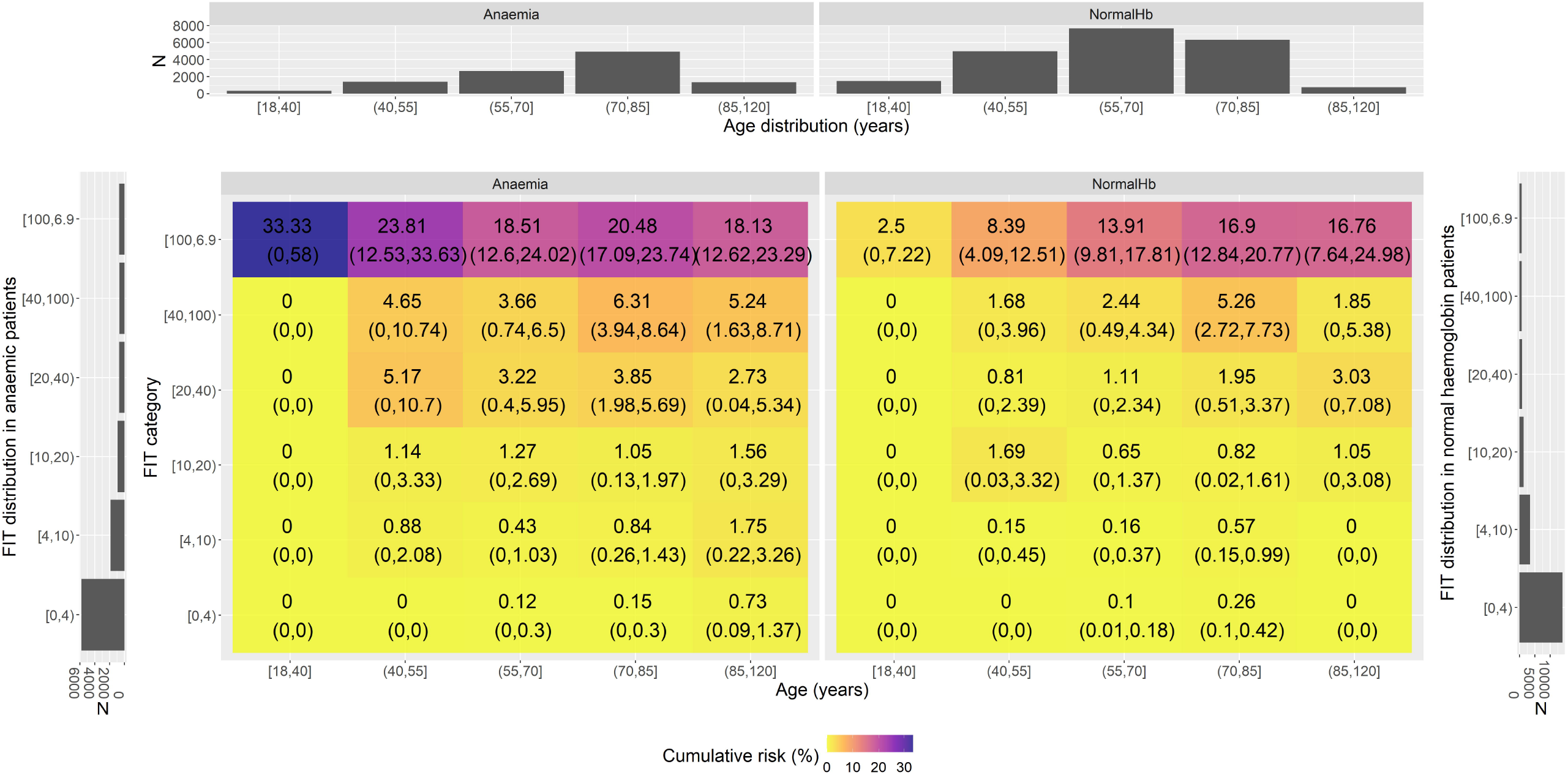
Heat map of CRC diagnoses by FIT level (5 categories), age group and anaemia (using only patients with blood tests, n = 33694)

### 1-year cumulative CRC risks by age, anaemia, thrombocytosis, and FIT level

In patients who are not anaemic and have a normal platelet count with a FIT between 40-100 μg Hb/g faeces, only those over the age of 70 years met the 3% threshold for investigation. All patients over the age of 40 years with a FIT≥100 μg Hb/g faeces met the threshold for investigation (Figure 4). Using a lower 95% confidence interval bound of the Kaplan Meier estimate would require a FIT cut off of ≥40 μg Hb/g faeces for all patients with normal blood tests under 70 years, and ≥20 μg Hb/g faeces for patients over 85 years. In patients with abnormal platelets and anaemia the threshold for investigation at 3% was met at a younger age and lower FIT category for example a FIT between 10-20 μg Hb/g faeces and age of 40-55 years.

**Figure 4.**
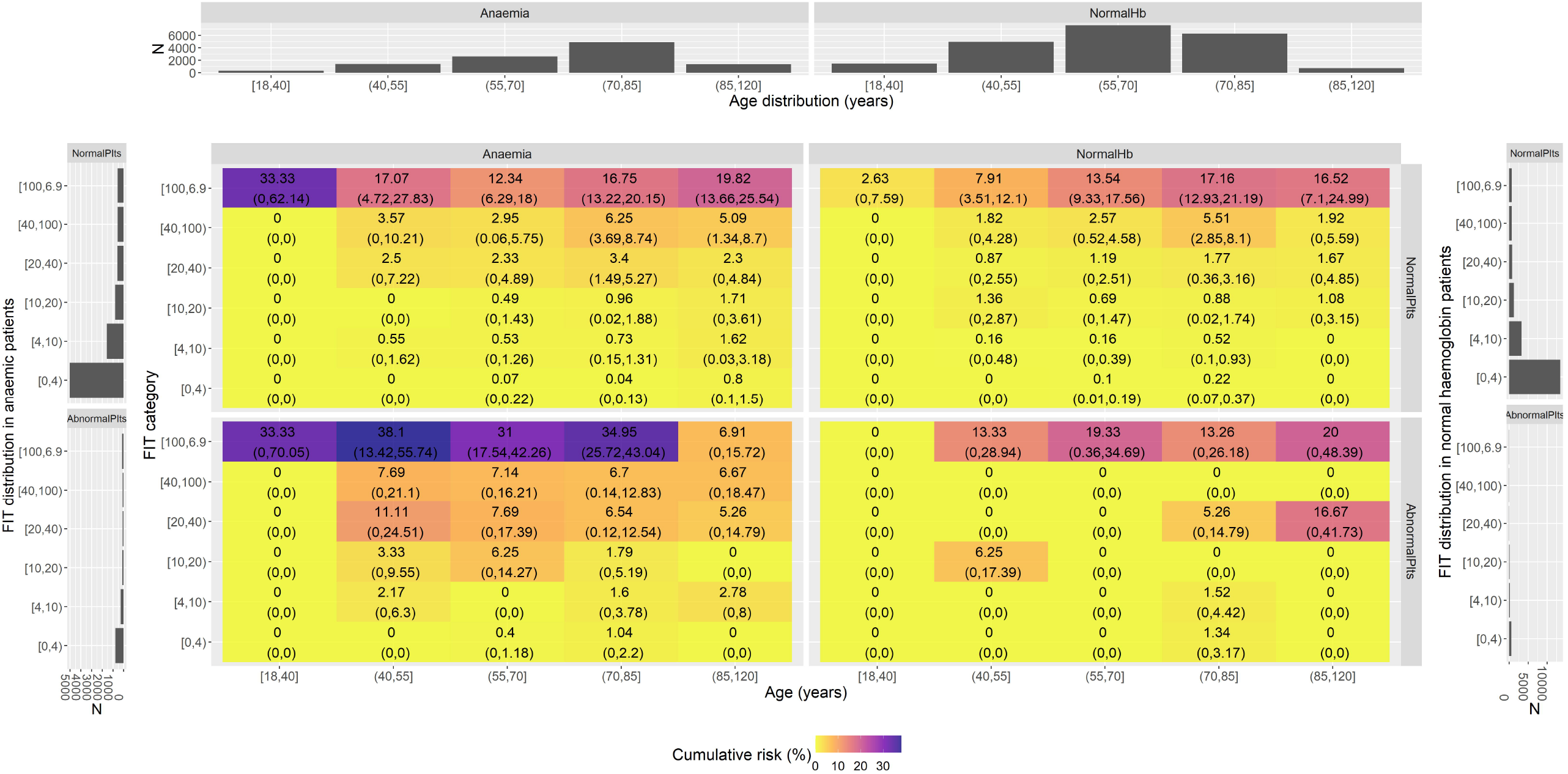
Heat map of CRC diagnoses by FIT level (5 categories), age group, thrombocytosis and anaemia (using only patients with blood tests, n = 33694).

### Estimates of investigations that could be re-purposed and potential cancers missed

7637 patients had a FIT test of ≥10 μg Hb/g faeces and so would have required a luminal investigation under a uniform binary cut off (as currently recommended nationally), with 53 CRCs missed in patients with FIT <10 μg Hb/g faeces. Table 3 shows the effect of selecting the FIT threshold that strictly meets the 3% threshold from the above heat maps, by showing the number of patients in those strata with a FIT value below the 3% 1-year risk of CRC threshold, but with a value of ≥10 μg Hb/g faeces (i.e. would have had a colonoscopy/CT colonography under a uniform FIT ≥10 μg Hb/g faeces cut-off). This showed that almost 1000 colonoscopies could be avoided by just using a FIT threshold of 100 μg Hb/g faeces for patients under 55, but an additional 13 cancers would be missed. Adding in anaemia almost halved the number of missed cancers in these strata but required delivery of 200 more urgent colonoscopies or equivalent. Abnormal platelets had a minimal additional change in the numbers after anaemia had been included. Using a 3% CRC threshold in low risk patients < 55 years for investigation including FIT, age and anaemia strata approximately 160-220 colonoscopies per 10000 FIT tests could be avoided at a cost of missing 1-2 CRCs (range indicating mean and lower 95% CI excluding 3% threshold).

**Table 3.**
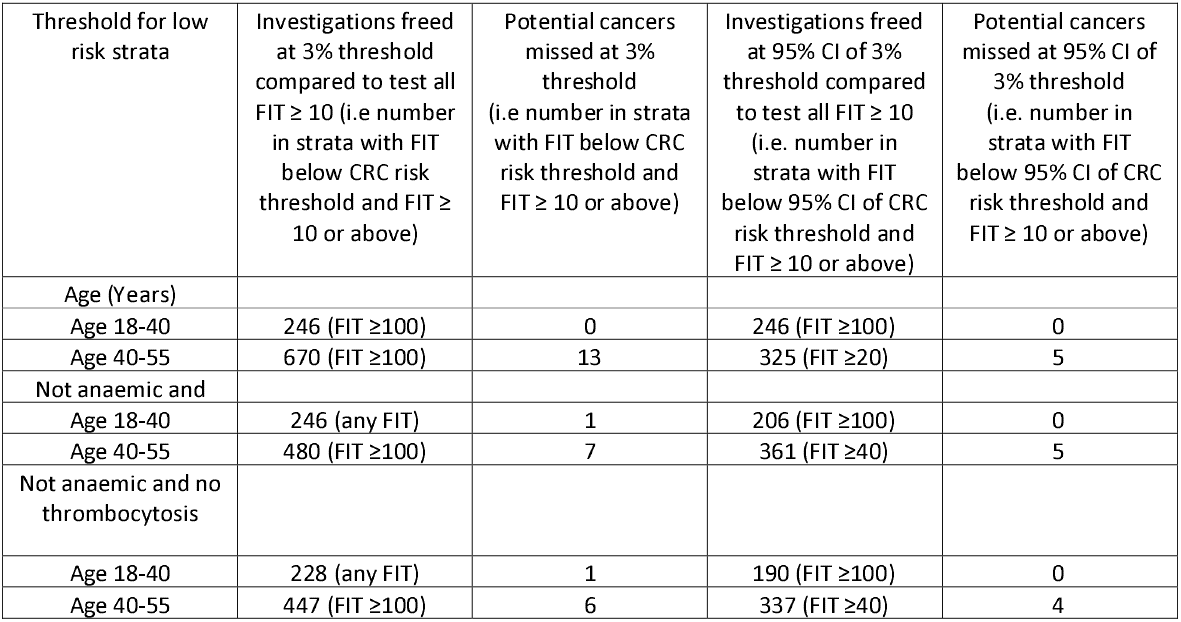
Estimated numbers of investigations that could be re-purposed (freed), and numbers of cancers potentially missed (using only patients and a single FIT test with blood tests, n = 33694).

## Discussion

We show that introduction of FIT in primary care as a gateway test to urgent pathways for CRC diagnosis, identified a population who have an overall 1-year cumulative risk of CRC of 1.5% following a FIT. We show that the CRC risk in FIT strata varies hugely by age and whether a patient is either anaemic or has thrombocytosis and, due to the large, representative nature of our study, we can give estimates of these differences. For example, non-anaemic patients do not meet the 3% threshold set by NICE for investigation until they have a FIT ≥40 μg Hb/g faeces. In contrast, those patients with anaemia meet the 3% threshold at a FIT of ≥20 μg Hb/g faeces. Patients under 40 years of age only meet the 3% threshold for investigation in those who have a FIT ≥100 μg Hb/g faeces and are anaemic. Estimating the risk in our study just above the current guideline recommended approach of a single cut-off at ≥10 μg Hb/g faeces, showed that patients with a FIT test of 10-20 μg Hb/g faeces had an overall uncensored risk of 25/2266 = 1.1%, well below the 3% threshold.

In our population we conservatively estimate that by using a stratified approach to FIT cut offs in low risk patients (<55 years and not anaemic) up to 600 urgent investigations (of which the majority are colonoscopy) for CRC can be forgone over 4 years within our catchment of over 37000 FIT tests at a cost of missing approximately 4 CRCs. When extrapolated to the whole country this could represent a re-purposing of 160-220 colonoscopies per 10000 FIT tests carried out at a cost of 1-2 cancers missed (using lower to mid-point estimates) for other purposes such as screening, surveillance or routine investigation. This compares to the current nationally recommended approach (FIT>10 μg Hb/g) which leads to over 2000 investigations per 10000 FIT tests. Our stratified results show that using more information from blood tests, and varying the FIT cut off can change the balance between the number of tests performed and the number of cancers missed in the investigation of symptomatic patients for CRC. The balance of investigations required, cancers diagnosed and missed is crucial to consider together when attempting to optimise diagnostic accuracy and health service provision in the real world. Consensus among all stakeholders needs to be reached on the threshold (risk of CRC) at which urgent investigation should be triggered, taking all these factors into account to optimally define this balance. This approach might allow the released diagnostic capacity to be used to support the lowering of the screening age in CRC screening in line with the NHS Long Term Plan^12^.

### Strength and Limitations

The size of the cohort and number of cancers diagnosed means there is sufficient power to stratify our results to understand the additional benefits of using age, sex and blood tests to identify those patients most at risk of CRC and potentially in which groups investigation can be safely avoided. It is important to note that these data reflect FIT in clinical use but still mirror the findings of research studies on FIT in selected populations. We used the OC-Sensor™ platform (Eiken Chemical Co., Tokyo, Japan) to determine FIT levels. Other analysers are utilised in symptomatic pathways with the other most frequently used being the HM-Jackarc analyser. The analysis presented focuses on the diagnosis of CRC. Other diagnoses such as inflammatory bowel disease (IBD) and polyps need to be considered as FIT may present opportunities to diagnose IBD earlier or treat potentially premalignant polyps however for the context of this study we have focused on the diagnosis of CRC as this is the primary purpose of the urgent diagnostic cancer pathways. However, freeing up diagnostic capacity could allow these patients more timely access to diagnosis. Finally, it is important to note the relatively high risk of death from other causes in our cohort which means that previously reported risks of CRC from studies that did not account for loss to follow up, or the competing risk of death will be overestimates of the 1-year cumulative risk of CRC. The high non-CRC death risk will potentially have impacted studies of diagnostic accuracy also where follow up was at least 1-year.

Our study assessed existing empirical categorisations of FIT, age and anaemia. Ideally, further optimisation and validation of pathways could be achieved by deriving and validating cut offs and strata using continuous modelling of FIT, age and blood test results in this and other population based datasets.

### Context of what is already known

A recent review of 28 832 patients^19^, selected for urgent referral and fully investigated, found a pooled sensitivity of 88.7% and specificity 80.5% using a cut off of 10 μg Hb/g faeces for CRC. In a further systematic review (16 studies, *n* = 35,945) the summary estimates of sensitivity and specificity were 91.0% (95% CI: 88.9, 92.7) and 75.2% (95% CI: 69.6, 80.1) for all patients with a cut off 10 μg Hb/g faeces for CRC. Furthermore a systematic review of FIT testing in primary care reported the at a cut off of 20 μg Hb/g faeces only one additional CRC would be missed per 1000 patients investigated at a CRC prevalence of 2%^20^. A number of other research studies have observed optimal FIT cut offs of ≥20 μg Hb/g faeces or higher^11 18^. NICE recommended a threshold for investigation of patients suspected of having CRC of 3% yet current studies often have a detection risk of just 1-2% suggesting further optimisation of the pathway could be achieved to identify patients above a 3% threshold of CRC^7 21^. A 3% threshold for investigation would result in fewer investigations such as colonoscopy which currently have limited capacity due to COVID related backlogs and increasing demands for investigation^5^. However, this need to reduce investigations has to be set against the need to safety net and minimise the likelihood of missed cancer diagnoses to ensure the successful implementation of symptomatic FIT pathways.

Higher cut offs for FIT above the currently recommended ≥10 μg Hb/g faeces cut off have been previously suggested. For example, an optimal FIT cut off of 19 μg Hb/g faeces was found in 5040 patients giving a sensitivity of 85.4% in a population with a risk of cancer of 3.0% (151/5040)^11^. The authors suggest a tailored approach to the use of FIT and produced estimates for optimal cut offs based on age and referring symptoms but focused on diagnostic accuracy rather than, as we do, the balance of risks and benefits to the health service as a whole. In an analysis of a population from Oxford, UK of 16604 patients with low risk symptoms for CRC and a CRC risk of 0.8% the addition of blood tests to FIT test results whilst improving specificity decreased sensitivity for the diagnosis of CRC^21^. The authors concluded FIT plus bloods tests did not improve discrimination for CRC however this was a low-risk population, and the authors did not consider the threshold at which investigation should be performed. Our data on the additional value of blood tests at the lower end of the fHb spectrum is consistent with other studies, including two from Scotland that have recently reported ^22 23^. Data from Tayside, the first to describe FIT for symptomatic patients in the UK, has also shown the potential value of haemoglobin and microcytosis in optimising triage^23^. Our results suggest that FIT, blood tests, and age could be used to refine protocols to implement FIT into 2WW pathways maintaining a balance between detection of CRC and the need to undertake endoscopic investigations.

Previous attempts to optimise FIT with other markers have shown limited benefit (FAST, COLONPREDICT)^24 25^. However, these tools have used different combinations across the full range of fHb results – it is unlikely that any marker will add to the value of high fHb (≥100 μg Hb/g faeces or similar). Reduction of missed CRC below any threshold for urgent referral, based on a FIT result alone or in combination with other markers, may be improved by repeat testing)^26 27^. We have not included the repeat tested group in our analysis. Although further work is required to validate this approach the “cost” – financial and otherwise of a missed CRC is much higher than that of a repeat FIT test. As such, the two approaches may be complementary in improving the use of FIT. Optimisation of FIT is likely to be of greatest value when the fHb result is “intermediate”, although a consensus definitions of such terms is required^28^.

### Clinical Significance

We show the 1-year cumulative risk of CRC among people who are referred for symptoms and carry out a FIT test varies hugely depending on the levels of FIT, their age and whether they have either anaemia and/or thrombocytosis. These figures are likely to be generalisable to the whole of England and most of the UK where FIT is being widely used to triage patients for investigation. Furthermore, our work demonstrates that the current FIT cut off recommended by NICE and others leads to populations that have a 1-year cumulative risk of CRC much lower than anticipated. This inevitably contributes to the overwhelming backlog for relevant investigations, in particular colonoscopy. If a nationally agreed cancer threshold of 3% is to be applied then current guidance may need to reconsider what FIT cut offs should be recommended, and how age and anaemia should be included in these pathways. Operational delivery of such pathways is possible as evidenced by the uptake of FIT usage in our population.

## Data Availability

This work uses data that has been provided by patients and collected by the NHS as part of their care and support. Under the Data Protection Impact Assessment approval for this work (DPIA reference: IG0889) we are unable to share the original data outside Nottingham University Hospitals NHS Trust.

## Ethics approval statement

HRA and Health and Care Research Wales (HCRW) approval was given for this study - IRAS project ID: 312362; Protocol number: 22ON007; REC reference: 22/HRA/2125; Sponsor: Nottingham University Hospitals NHS Trust

## Patient and public involvement

There was no public or patients involved in this work.

## Authorship statement

All authors were part of the conceptualisation of the work. JW, DJH and CJC wrote the applications for permission to access the data, the protocol for the study and gained HRA approval. JJ carried out data management with CJC, and CJC carried out the analysis. DJH, JW and CJC wrote the first draft of the manuscript, and all authors were involved in the writing, reviewing and editing drafts of the paper and approving the manuscript for submission. DJH and CJC are guarantors.

