## Supplementary material for "Assessing empirical thresholds for investigation in people referred on a symptomatic colorectal cancer pathway: a cohort study utilising faecal immunochemical and blood tests in England"

Supplementary figure 1. The current Nottingham Rapid Colorectal Cancer Diagnosis pathway since modification in November 2021 and summary of changes since introduction. *advised to undertake PR if not already performed

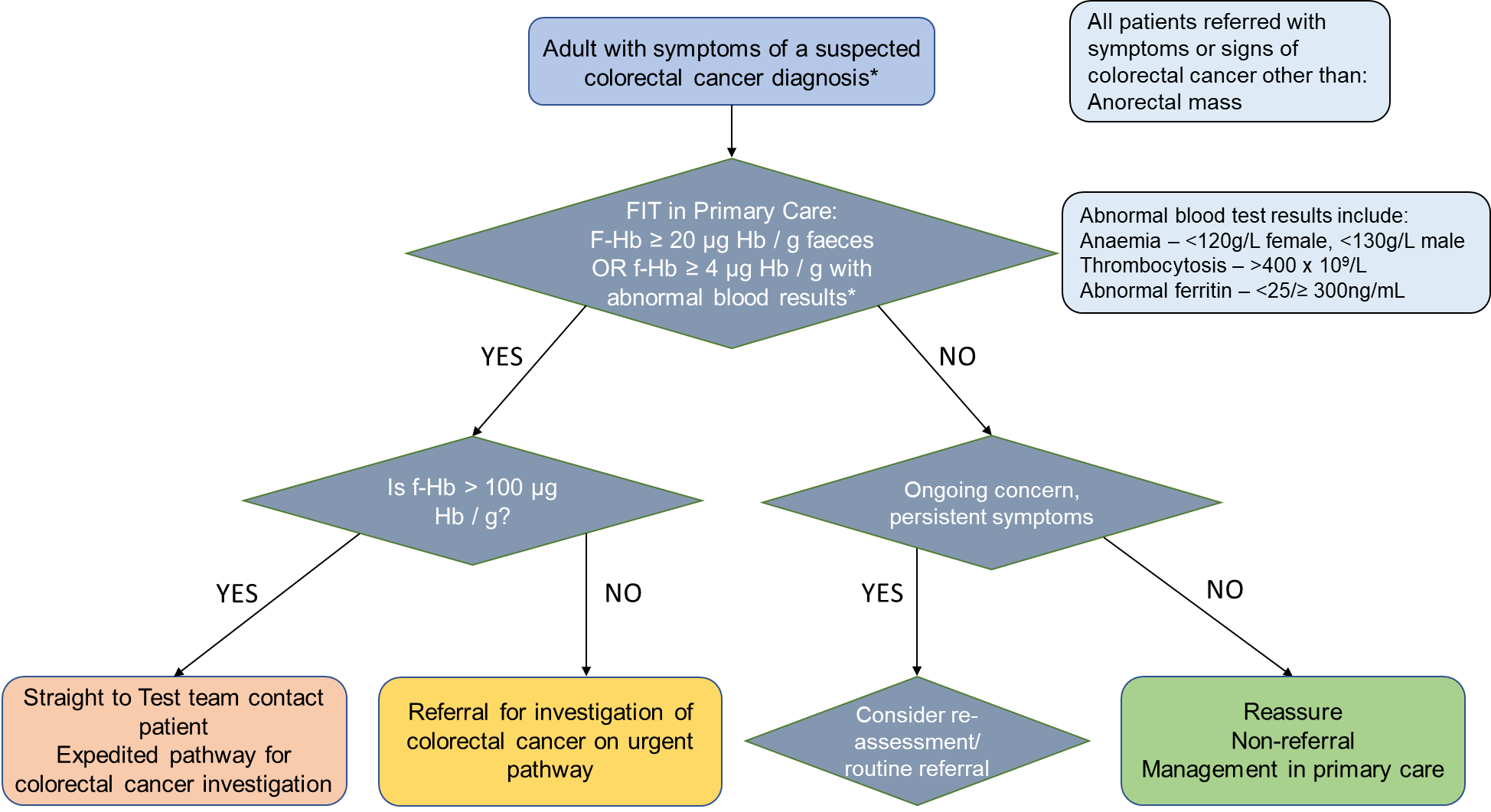

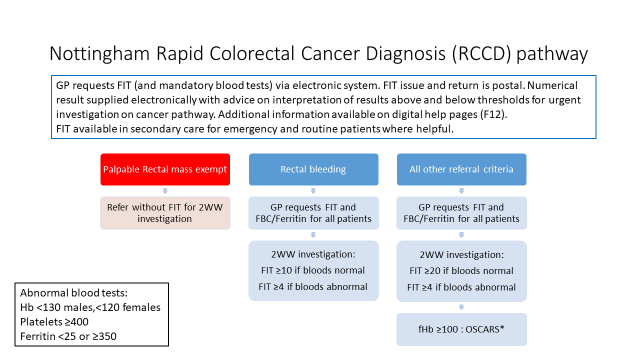

OSCARS – One stop Surgical assessment, Colonoscopy and Radiological Staging – dedicated colonoscopy lists with senior surgeons and protected slots for radiology when required.

Supplementary figure 2. Flow chart of included people and FIT tests

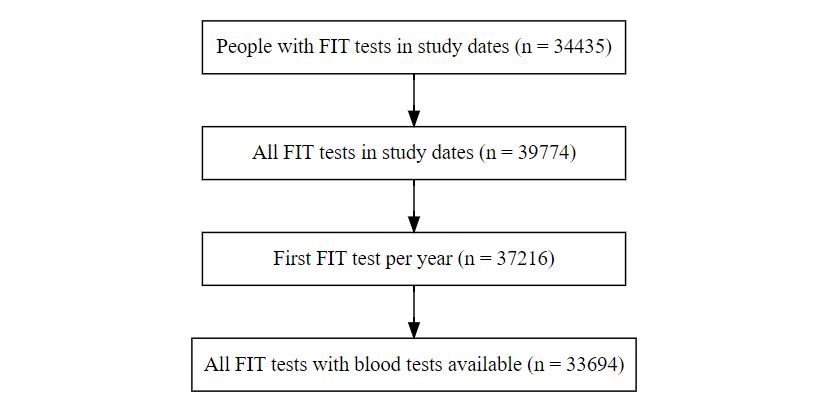

**Supplementary results**

The competing risk of non-CRC death is more likely than CRC (non-colorectal cancer death 4.8% versus colorectal cancer diagnosis 1.5%), and this differential censoring occurs throughout the follow up with a difference of 18 days between median time to censoring.

Supplementary table 1. Median days to non CRC

| **FIT ≥10** | **Median days to non CRC death** | **(min, IQR, max)** |
| --- | --- | --- |
| TRUE | 176 | (11, 91, 266, 365) |
| FALSE | 158 | (3, 94, 268, 365) |

Supplementary figure 3. Counts of non-colorectal cancer death by time since FIT test, by FIT result category

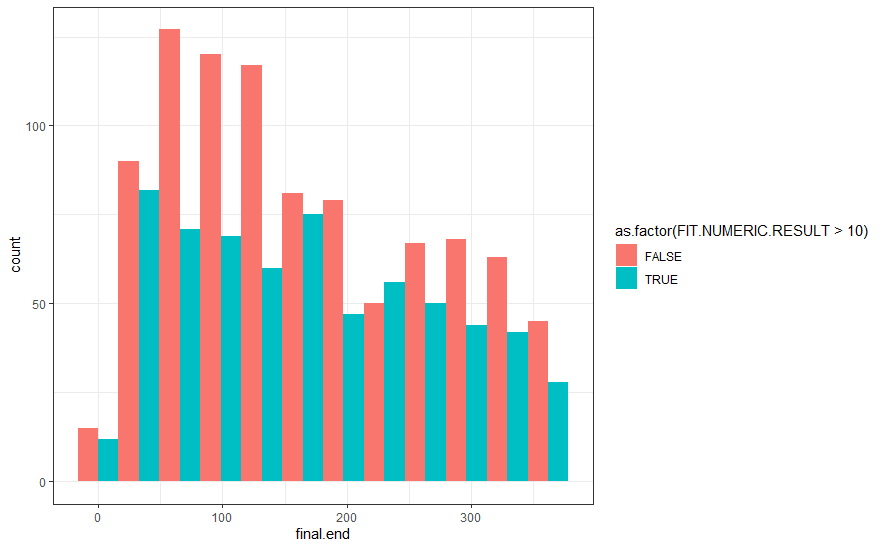

Supplementary figure 4. Histogram of non-colorectal cancer death by time from FIT test, by FIT result category

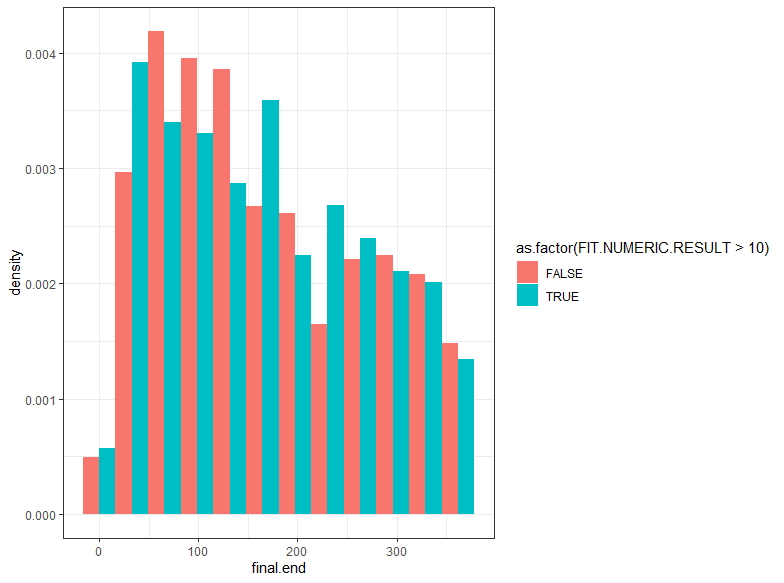

There is approximately a 30 day median delay in diagnosis in the FIT <10 group, however, most colorectal cancers in both groups are diagnosed early in the one year follow up.

Supplementary table 2. Median days to colorectal cancer diagnosis from FIT test.

| **FIT ≥10** | **Median days to colorectal cancer diagnosis** | **(min, IQR, max)** |
| --- | --- | --- |
| TRUE | 33 | (10, 24, 56, 358) |
| FALSE | 66 | (15, 38, 132, 320) |

Supplementary figure 5. Counts of colorectal cancers by time from FIT test, by FIT result category

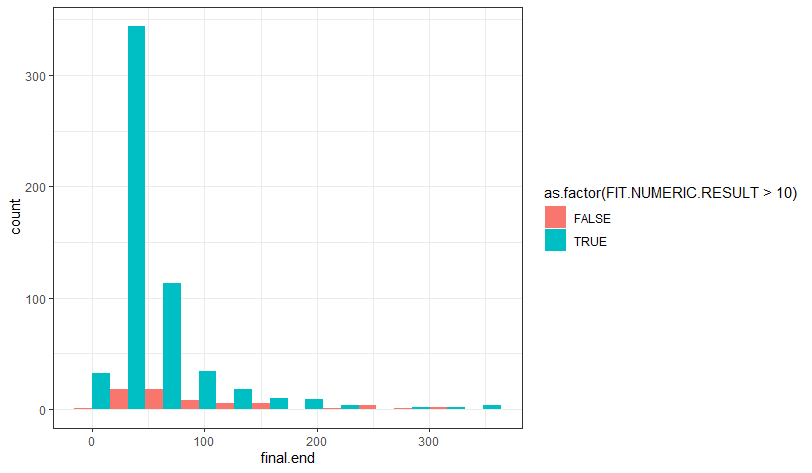
 Supplementary figure 6. Histogram of colorectal cancers by time since FIT test, by FIT result category

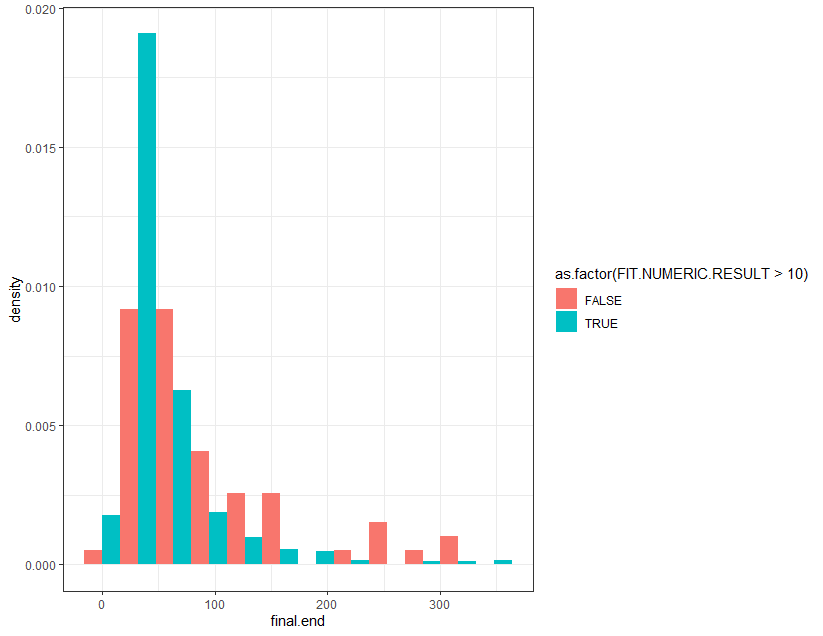
